# Epidemiological and Clinical Characteristics of Pediatric Intoxications: A Retrospective Study

**DOI:** 10.64898/2025.12.13.25342187

**Authors:** Mohamed Boulahia

**Affiliations:** Faculty of Medicine, University of Algiers, Algeria

**Keywords:** Poisoning, Child, Caustics, Carbon Monoxide Poisoning, Retrospective Studies, Algeria

## Abstract

**Background:** Pediatric poisonings are a significant cause of emergency admissions, often linked to accessible household toxins. This study investigates the epidemiological patterns, clinical presentations, and management strategies at a tertiary care hospital in Algiers, Algeria.

**Methods:** A retrospective descriptive analysis was conducted on anonymized pediatric poisonings cases. Variables included age, sex, type of ingestion, intent (accidental vs. voluntary), substances involved, endoscopic findings, hospitalization duration, and management approach.

**Results:** 59 pediatric poisoning cases were analyzed, which were predominantly accidental and affected children aged 0–6 years. Medication (n=24) and caustic ingestions (n=25) were the most frequent agents. Caustic cases, which primarily affected older toddlers, led to moderate mucosal injury in three patients. Carbon monoxide (n=8) affected a wide age range, including infants and adolescents. Management was mainly supportive; while most medication cases were discharged in two days, caustic cases required hospitalization for up to 14 days. These findings highlight the need for targeted public health interventions.

**Conclusion:** Pediatric poisonings in this setting are largely preventable. Unsafe storage of medications and caustic agents is the primary contributor. Public health interventions focusing on household safety, caregiver education, and standardized clinical protocols are essential.

**Key Points:** *What is known:*

- Pediatric poisonings are common emergency admissions.
- Toddlers are the most vulnerable age group.
- Unsafe household storage is a major risk factor.

*What is added:*

- First detailed Algerian hospital-based analysis of pediatric intoxications.
- Characterizes the dual burden of caustic and medication poisonings in a North African pediatric setting, with caustic ingestions accounting for the most prolonged hospitalizations and requiring endoscopic evaluation.
- Demonstrates that carbon monoxide poisoning affects a broad age range including infants, highlighting environmental domestic exposure as a distinct and underappreciated risk in this context.

## Introduction

Pediatric poisonings represent a significant global health challenge and remain among the leading causes of emergency department visits in children, particularly those under five years of age [1,2]. Most intoxications result from unintentional exposures, often linked to inadequate storage of medications and household chemicals, limited caregiver awareness, and the widespread availability of toxic substances within domestic environments [3]. The clinical spectrum ranges from mild, self-limiting symptoms to severe, life-threatening conditions requiring urgent intervention.

Epidemiological patterns of pediatric intoxications vary across regions, influenced by socioeconomic factors, cultural practices, and healthcare infrastructure [4,5]. In Algeria, comprehensive data on the burden, clinical outcomes, and management of pediatric poisonings remain scarce, limiting the development of targeted prevention and intervention strategies [6].

This study aims to address this gap by providing a detailed retrospective analysis of pediatric intoxications presenting to a tertiary care hospital in Algiers, Algeria. The focus is on epidemiological characteristics, clinical presentations, and management outcomes. The findings are intended to inform local public health policies and clinical protocols, thereby contributing to improved prevention and care of pediatric intoxications in Algeria.

## Materials & Methods

### Study Design and Setting

This retrospective descriptive study was conducted in the Department of Pediatrics, Faculty of Medicine, University of Algiers, between January 2024 and June 2025. The department serves a diverse urban population, providing emergency and inpatient pediatric care.

### Population

A total of 59 pediatric poisoning cases were identified during the study period and included in the analysis. Patient data were anonymized to ensure confidentiality and compliance with institutional and ethical guidelines.

### Inclusion criteria

Children between the ages of 0 and 15 years who presented to the pediatric emergency department with a confirmed diagnosis of accidental or intentional poisoning were eligible for inclusion. Confirmation was based on clinical history provided by caregivers, direct observation of ingestion or exposure, and/or laboratory evidence consistent with toxic substance exposure. Only cases with complete medical records, including demographic data, clinical presentation, management, and outcome, were included to ensure reliability of analysis.

### Exclusion criteria

Cases were excluded if medical documentation was incomplete, if the diagnosis of poisoning remained uncertain after clinical and laboratory evaluation, or if the child had been transferred from another hospital without comprehensive clinical data. Patients with chronic intoxication or poisoning secondary to underlying metabolic disorders were also excluded to maintain homogeneity of the study population.

### Data Collection

Data were systematically extracted from hospital medical records and included demographic variables (age, sex), intoxication categories, specific toxic agents involved, intent of ingestion (voluntary vs. accidental), clinical findings including endoscopic evaluations (Esophagogastroduodenoscopy [EGD]), hospitalization duration, management strategies, and patient outcomes. Data accuracy was ensured through cross-verification by clinical staff.

### Ethical Considerations

This study was conducted in accordance with the Declaration of Helsinki. Given its retrospective design involving exclusively anonymized data and no direct patient intervention, formal approval by the institutional ethics committee of the Faculty of Medicine, University of Algiers was not required. Patient confidentiality was maintained throughout data collection and analysis.

### Outcomes

#### Primary outcome

The primary outcome measure was the clinical course and recovery status of poisoned children. This included evaluation of symptom progression, requirement for intensive care unit (ICU) admission, and survival status at discharge. Recovery was defined as resolution of acute symptoms without long-term sequelae, whereas adverse outcomes encompassed prolonged hospitalization, ICU stay, or death.

#### Secondary outcomes

Secondary outcomes were designed to capture the broader epidemiological and clinical profile of pediatric poisoning. These included the type of toxic agent involved such as pharmaceuticals, household chemicals, hydrocarbons, plants, or other substances, the route of exposure, whether oral ingestion, inhalation, dermal contact, or mixed, and the time interval between exposure and presentation to the emergency department. Additional measures comprised the occurrence of complications, including neurological, respiratory, gastrointestinal, or cardiovascular events during hospitalization, as well as the length of hospital stay, recorded in days from admission to discharge. Collectively, these outcomes provided a comprehensive assessment of both the immediate clinical impact and the overall resource utilization associated with pediatric poisoning cases.

## Results

The results of this study consolidate the epidemiological, clinical, and hospitalization data for pediatric poisonings at a tertiary care hospital in Algiers, Algeria. The majority of cases were accidental and predominantly involved children aged 0 to 3 years. Medication and household products were the most common agents implicated.

### Epidemiological Profile

A total of 59 pediatric poisoning cases were analyzed. The majority were accidental (56/59, 94.9%), while only three cases (5.1%) were intentional; all intentional cases involved medication. Children aged 0-3 years were most frequently affected (35/59, 59.3%), followed by those aged 4-10 years (15/59, 25.4%) and adolescents aged 11-15 years (9/59, 15.3%). Overall, males accounted for 32 cases (54.2%) and females for 27 cases (45.8%).

### Distribution by Toxic Agent

The 59 cases analyzed included 24 medication poisonings, 25 ingestions of caustics and household products, 8 carbon monoxide poisonings, 1 electrical injury, and 1 plant ingestion. Medication poisonings involved agents such as paracetamol, neuroleptics, and antihypertensives, with an equal distribution between females and males.

The category of caustics and household products was more frequent in males and included a mix of corrosive agents and domestic toxins. Specific agents included drain openers (6), spirit of salt (Hydrochloric Acid) (2), and formalin (2), which pose direct risks of mucosal injury. Additionally, this group included non-corrosive household toxins such as rat poison (rodenticides) (6), insecticides (3), eau de parfum (4), and thinner (2). Endoscopic evaluation (EGD) was reserved for suspected corrosive ingestions; it was performed in seven cases, revealing grade IIA esophagitis and gastritis lesions in three patients, indicating moderate mucosal injury.

CO poisonings affected a broad age range: adolescents (14-15 years) and younger children, including infants (10 months and 19 months). This highlights that while adolescents were frequently affected, infants are also at risk due to domestic environmental exposures.

### Clinical Management

Medication poisonings (n=24) were generally mild: 16 patients (66.7%) were discharged after short observation, seven (29.2%) underwent gastric lavage, and one (4.1%) required antibiotic therapy for secondary complications.

Ingestions of caustics and household products (n=25) demanded more intensive care: 12 patients (48%) received combined therapy with antibiotics, proton pump inhibitors (PPI), and cessation of oral feeding; two (8%) required gastric lavage (performed only for non-corrosive agents within this category); and 11 (44%) were discharged after observation.

Carbon monoxide cases (n=8) were managed with supportive care and oxygen therapy, with all patients recovering fully without prolonged hospitalization. The single cases of electrical injury and plant ingestion were treated conservatively with observation and supportive care, both resulting in uncomplicated discharge.

### Hospitalization Outcomes

Hospitalization durations were generally short, with most patients discharged within two days. Medication poisonings typically required 1-4 days (J0-J4), while caustic and household ingestions extended up to 14 days (J0-J14). Carbon monoxide, electricity, and plant exposures did not require prolonged admission, as patients were discharged directly after supportive management. Overall, 53/59 patients (89.8%) were discharged without complications, while six (10.2%) required more intensive therapy.

An overview of the study’s key findings regarding the distribution of cases, demographics, hospitalization duration, and outcomes is summarized in Table 1 and illustrated in Figure 1.

**Table 1.** Patterns and Outcomes of Pediatric Poisonings Distribution of pediatric poisoning cases by category, sex, age group, intent, hospitalization duration, and main outcomes. **Footnotes:** J0 = first day of admission; J4 = fourth hospital day; J14 = fourteenth hospital day. N/A = not applicable, patients discharged directly after supportive management.

| Category | Cases | Female | Male | Age 0–3<br>years | Age 4–10<br>years | Age 11–<br>15 years | Voluntary | Accidental | Hospitalization<br>(avg days) | Outcome<br>(main) |
| --- | --- | --- | --- | --- | --- | --- | --- | --- | --- | --- |
| Medication | 24 | 12 | 12 | 14 | 6 | 4 | 3 | 21 | J0–J4 | Mostly discharge |
| Caustics &<br>Household<br>Products | 25 | 10 | 15 | 16 | 6 | 3 | 0 | 25 | J0–J14 | Mixed discharge<br>& therapy |
| Carbon<br>monoxide | 8 | 5 | 3 | 3 | 2 | 3 | 0 | 8 | N/A | Discharge |
| Electricity | 1 | 0 | 1 | 1 | 0 | 0 | 0 | 1 | N/A | Discharge |
| Plants | 1 | 0 | 1 | 1 | 0 | 0 | 0 | 1 | N/A | Discharge |
| Total (n=59) | 59 | 27 | 32 | 35 | 14 | 10 | 3 | 56 | — | — |

**Figure 1.**
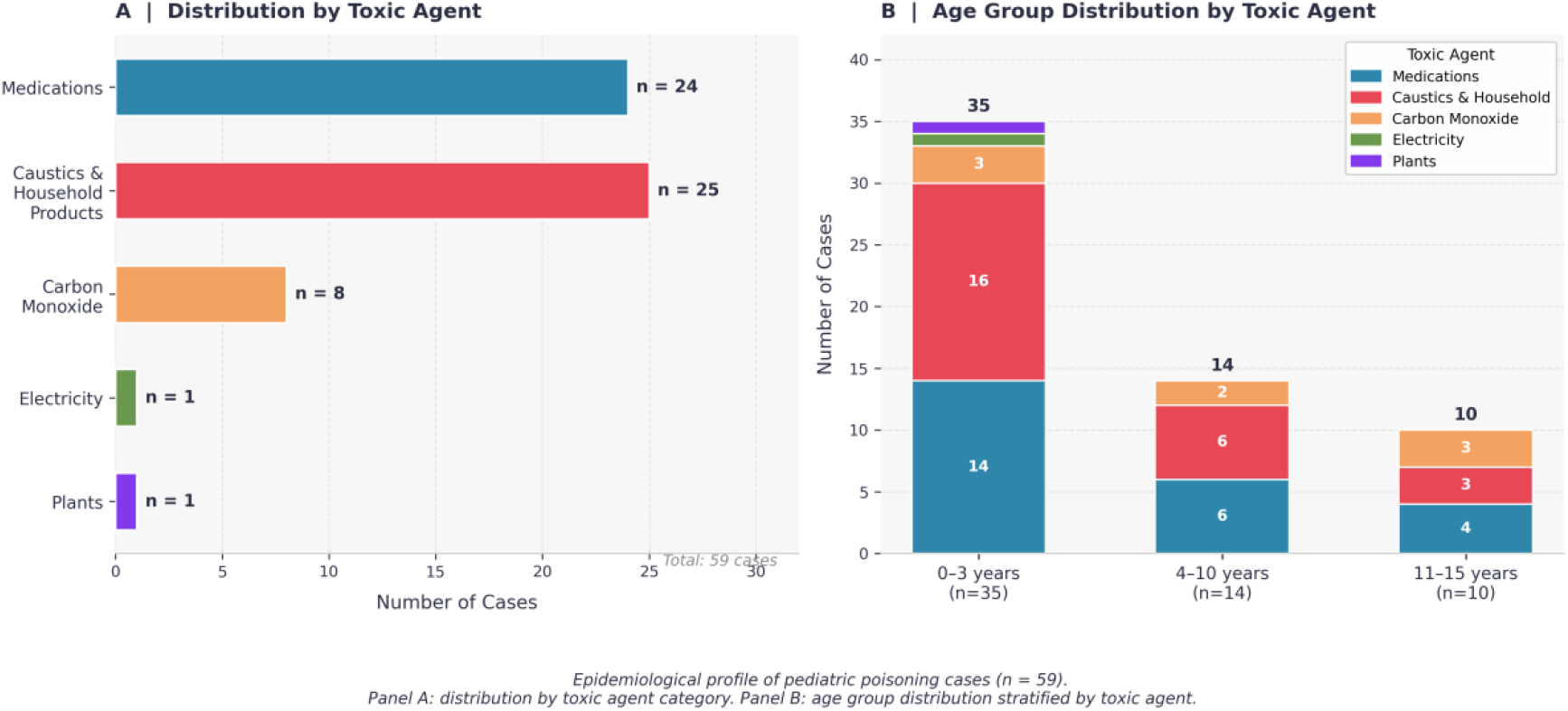
Epidemiological profile of pediatric poisoning cases (n = 59). Panel A: distribution by toxic agent category. Panel B: age group distribution stratified by toxic agent.

## Discussion

This study demonstrates that pediatric poisonings at a tertiary care hospital in Algiers, Algeria predominantly affect toddlers aged 1-3 years and are mostly accidental. These findings are consistent with global data showing that children under five are the most vulnerable age group due to exploratory behavior and easy access to household substances. Medication poisonings were common, with neuroleptics and antihypertensives frequently involved, underscoring the need for secure storage and child-resistant packaging [1,2]. Similar trends have been reported in India and South Africa, where pharmaceuticals accounted for a large proportion of pediatric intoxications [3,4].

Caustic ingestions posed significant risks due to potential long-term morbidity. Endoscopic evaluations revealed grade IIA lesions in multiple cases, highlighting the importance of early assessment and intervention [5,6]. Comparable findings were reported in Turkey, where caustic substances were among the most frequent agents and associated with prolonged hospitalization [7]. The wide range of caustic agents in our cohort reflects the accessibility of household chemicals, emphasizing the need for public health measures to limit exposure [8,9].

Carbon monoxide poisoning affected eight children, primarily older children and adolescents, but notably included two infants. This underscores that CO exposure is not limited to older age groups. Similar age patterns have been documented in Turkey and the United States, where environmental exposures placed both toddlers and adolescents at risk [10,11]. Preventive measures such as improved ventilation and installation of CO detectors remain critical [12].

Hospitalization durations were generally short, suggesting mild clinical presentations. However, the presence of severe cases requiring extended hospitalization highlights the need for continuous clinical vigilance and protocol optimization. Previous studies in the Middle East and Africa also reported short hospital stays for most cases, with longer admissions linked to caustic ingestions and severe intoxications [13,14].

Management strategies in our cohort highlight the predominance of supportive care in pediatric intoxications, with specific interventions tailored to the agent involved. Medication-related poisonings were generally mild, requiring only observation or gastric lavage, consistent with international guidelines [1,2]. Caustic ingestions demanded more aggressive management, including antibiotics, proton pump inhibitors, and temporary cessation of oral feeding [5,6]. Carbon monoxide poisonings were effectively managed with oxygen therapy [10,11]. Rare cases such as electrical injury and plant ingestion illustrate the diversity of pediatric poisonings but reinforce that supportive care remains the cornerstone of management across presentations [3,7].

### Limitations

This study has several limitations. First, it was conducted at a single center, which may limit generalizability. Second, the retrospective design relied on medical records, and incomplete documentation led to the exclusion of some cases. Third, laboratory confirmation was not available for all exposures, and outcomes were limited to hospitalization data without long-term follow-up. Despite these limitations, the study provides valuable insights into the epidemiological and clinical patterns of pediatric poisonings in Algeria.

## Conclusions

Pediatric poisonings in this Algerian hospital setting are largely preventable, with medications and caustic substances emerging as the most frequent agents and young children being disproportionately affected. The predominance of accidental ingestions in toddlers aged 0–3 years underscores the role of unsafe household storage and limited caregiver awareness as primary modifiable risk factors. These findings support the implementation of targeted preventive measures, including secure storage of pharmaceuticals and hazardous household chemicals, child-resistant packaging policies, and structured caregiver education programs on early recognition of poisoning signs. At the clinical level, standardized management protocols—particularly for caustic ingestions requiring endoscopic evaluation and extended hospitalization—are essential to minimize complications and optimize outcomes. Coordinated efforts among healthcare providers, public health authorities, and caregivers are necessary to reduce the burden of pediatric poisoning in this setting.

## Data Availability

The datasets generated and/or analyzed during the current study are no longer available, as the retrospective data collection was anonymized and not retained beyond the study period. All results derived from these data are fully reported within the manuscript.

## Acknowledgments

The author thanks the staff of the Department of Pediatrics, Hassen Badi Hospital for their support.

## Financing Sources

This research did not receive any specific grant from funding agencies in the public, commercial, or not-for-profit sectors.

